# Ecological Momentary Assessment of Anxiety and Depression in Individuals with Early-Treated Phenylketonuria

**DOI:** 10.64898/2026.10.23.26366355

**Authors:** Braden E. Hayse, Sean P. Lane, Shifali Singh, Stephanie Sacharow, Benjamin D. Goodlett, Shawn E. Christ

**Author notes:** Address correspondence to: Shawn E. Christ, Department of Psychological Sciences, University of Missouri, 210 McAlester Hall, Columbia, MO 65203.

## Abstract

**Background:** In addition to cognitive difficulties, individuals with early-treated phenylketonuria (PKU) may be at increased risk for anxiety and depression. However, previous findings are mixed, potentially due to reliance on retrospective self-report measures that are vulnerable to recall bias and lose resolution of day-to-day symptom fluctuations. The present study used ecological momentary assessment (EMA) to evaluate anxiety and depressive symptoms in adults with PKU in real-world settings.

**Methods:** A sample of 30 adults with early-treated PKU and a demographically-similar sample of 30 adults without PKU completed smartphone-based EMA assessments four times daily over seven consecutive days (*n_Total_*=1,384). Anxiety and depression symptoms and processing speed task performance were assessed at each timepoint. Previous week retrospective measures of mood symptoms were also collected following the EMA period.

**Results:** EMA compliance was high (82.4%). Momentary analyses revealed that PKU participants reported significantly greater anxiety and depression symptoms as well as poorer processing speed performance compared to healthy controls. In contrast, no significant group differences were identified using retrospective mood measures. Within the PKU group, Phe levels were not significantly associated with mood symptom severity or processing speed.

**Conclusions:** EMA detected PKU-related differences in psychological symptoms that were not identified by traditional retrospective assessments. These findings highlight the feasibility and sensitivity of EMA for characterizing everyday functioning in individuals with PKU. EMA may provide a more accurate assessment of symptom burden and may be particularly valuable for studying rare clinical populations, where intensive longitudinal data collection can maximize information gained from relatively small samples.

**Highlights:**

- EMA detected PKU-related elevations in anxiety and depression symptoms
- Group differences were not evident on retrospective mood measurement
- EMA may be particularly valuable for studying rare clinical populations like PKU

## 1. INTRODUCTION

Phenylketonuria (PKU) is an autosomal recessive disorder characterized by a deficiency in phenylalanine hydroxylase, an essential enzyme for the metabolism of the amino acid phenylalanine (Phe) to tyrosine. As a result, PKU is associated with higher-than-normal levels of Phe as well as decreased levels of tyrosine and its metabolites such as dopamine (Pardridge, 1998). Brain levels of large neutral amino acids (LNAAs) such as tyrosine and tryptophan as well as their metabolites (e.g., dopamine, serotonin) are further impacted by the competition between the excess Phe and LNAAs to cross the blood-brain barrier as well as Phe-mediated inhibition of tyrosine hydroxylase and tryptophan hydroxylase activities in brain (thus impeding the synthesis of dopamine and serotonin; Curtius et al., 1981; Harding et al., 2014; Winn et al., 2018). High Phe levels also directly interfere with processes such as myelination and protein synthesis (de Groot et al., 2010; Pardridge, 1998) and are associated with oxidative stress damage to lipid, protein, and DNA macromolecules related to impaired antioxidant levels (Sanayama et al., 2011; Schuck et al., 2015; Sirtori et al., 2005; Sitta et al., 2009).

Untreated PKU can result in severe intellectual and neurologic impairment (Paine, 1957). Fortunately, the severe consequences associated with untreated PKU can be largely mitigated with early identification and treatment implementation aimed at reducing Phe levels (Ashe et al., 2019; González et al., 2011). Individuals with early-treated PKU, however, are still at risk for neurologic, cognitive, and psychological problems. For example, abnormalities in white matter and gray matter structures are consistent neurologic findings (e.g., Anderson & Leuzzi, 2010; Christ et al., 2010; Muri et al., 2022). In addition, a slight decrease in overall intellectual ability along with impairments in specific domains including executive function, attention, and processing speed is frequently reported (Clocksin et al., 2023; Palermo et al., 2017).

A growing literature also points to a higher prevalence of mental health concerns in individuals with early-treated PKU across the lifespan when compared to the general population, with internalizing disorders such as anxiety (Bilder et al., 2013; Boland et al., 2024; Koch et al., 2002; Manti et al., 2016; Pietz et al., 1997; Smith et al., 1988; Waisbren et al., 2017; Weglage et al., 2000) and depression (Bilder et al., 2013; Jahja et al., 2017; Koch et al., 2002; Manti et al., 2016; Pietz et al., 1997; Trefz et al., 2019; Waisbren et al., 2017; Weglage et al., 2000) representing the most widely reported comorbidities. Additionally, studies have found that poorer metabolic control (as reflected by higher Phe levels) is associated with increased symptoms of anxiety and depression in individuals with early-treated PKU (Clocksin 2023 in JINS; Clacy et al., 2014; Jahja et al., 2017; ten Hoedt et al., 2011). However, findings across studies have not been entirely consistent. Some investigations have failed to identify elevated rates of anxiety and/or depression in this population (Brumm et al., 2004; Channon et al., 2005, 2007; Clacy et al., 2014; Palermo et al., 2020), while others have not found a significant association between metabolic control and psychological outcomes (Bilder et al., 2013; Brumm et al., 2004; Palermo et al., 2020; Pietz et al., 1997; Ris et al., 1997; Weglage et al., 2000).

Importantly, the majority of these past studies relied on traditional self-report questionnaires or interviews. Such methods often require participants to evaluate and aggregate their past experience of symptoms over an extended time period (e.g., the past two weeks). This cumulative task is made even more difficult by the fact that anxiety and depression represent dynamic processes, the manifestation and experience of which fluctuate over time as a result of internal and external influences (Trull & Ebner-Priemer, 2020). In addition, such traditional methods are susceptible to recall bias (e.g., “peak-end” rule whereby intense moments and final moments heavily influence our recollection of events) and may be particularly problematic for clinical populations such as early-treated PKU with possible memory and/or introspection difficulties.

In contrast to retrospective assessment methods, ecological momentary assessment (EMA) represents a newer methodologic approach that involves repeated sampling of participants’ thoughts and experiences in real-time (aka “in the moment”) within their natural environments (Shiffman et al., 2008; Trull & Ebner-Priemer, 2020). By targeting momentary experiences, EMA can minimize recall bias and memory recollection demands. In addition, with the collection of data at multiple timepoints, it allows researchers to examine not only aggregate measures (e.g., average anxiety score) but also other facets of the data (e.g., intra-individual variability in anxiety scores over time) that would not otherwise be possible with more traditional single-timepoint methods. Lastly, by collecting real-time data within an individuals’ everyday life, EMA provides improved external and ecological validity over more traditional measures (Trull & Ebner-Priemer, 2014, 2020).

To date, EMA has been used to study a wide range of behaviors and psychological constructs in numerous clinical populations across the lifespan including individuals with anxiety (Farmer & Kashdan, 2014; Kashdan et al., 2013; Lamers et al., 2018), depression (Connolly & Alloy, 2018; Hepp et al., 2017; Thompson et al., 2012), attention deficit hyperactivity disorder (Skirrow et al., 2014), schizophrenia (Moran et al., 2017, 2018), autism (Khor et al., 2014), substance abuse (Mallard et al., 2018; Simpson et al., 2014), and eating disorders (Berg et al., 2013; Engel et al., 2013). It is also being incorporated into a growing number of clinical trials to evaluate the efficacy of behavioral and pharmacological treatments (for review & additional discussion, see Mofsen et al., 2019).

The present study aims to demonstrate the value and feasibility of EMA methodology for the study of psychological functioning, particularly anxiety and depression, in individuals with early-treated PKU. To this end, we used a cell phone-based application to collect real-time symptom data multiple times per day over the course of seven consecutive days in a sample of 30 adults with early-treated PKU and a demographically-similar sample of 30 healthy adults without PKU. We hypothesized that the early-treated PKU group would exhibit higher anxiety and depression symptom scores than the non-PKU group and that these differences would be more evident when assessed using EMA compared with a traditional single retrospective report. We also anticipated that, relative to the non-PKU group, the early-treated PKU group would demonstrate poorer performance on the processing speed task, as reflected by slower response times. Furthermore, we expected that such effects would be most pronounced in individuals with poorer metabolic control (i.e., higher Phe levels). In addition, we leveraged the richness of the dataset to explore potential associations between the outcome variables and temporal factors (e.g., time of day and day of the week). Finally, we examined the between-person and within-person reliability of the EMA measures in light of the novelty of this methodology for investigation among PKU individuals.

## 2. METHODS

### 2.1. Participants

A sample of 30 adults with early-treated PKU (18–40 yrs. of age) and a demographically-similar comparison group of 30 healthy adults without PKU participated in the study. While compliance with the EMA prompts was fairly high across both groups (82.8%), four participants (two PKU, two non-PKU) completed fewer than 60% of administered prompts and thus were excluded from further analysis. Demographic information for the remaining sample of 56 participants is presented in Table 1. There were no significant group differences in terms of age, sex, or years of education [*t*(54) < 1.22, *p* > .22, in all instances; χ^2^(1) = 0, *p* = 1 for sex].

**TABLE 1.** Demographic Information.

|  | PKU (*n* = 28) |  | Non-PKU (*n* = 28) |  | *p* |
| --- | --- | --- | --- | --- | --- |
|  | *M (SD)* | Range | *M (SD)* | Range |  |
| Age (years) | 30.1 (5.9) | 18 - 40 | 29.2 (6.2) | 18 - 41 | 0.59 |
| Sex (male/female) | 10/18 |  | 9/19 |  | - |
| Education (years) | 15.1 (2.5) | 8 - 18 | 15.9 (2.7) | 11 - 24 | 0.23 |
| Phenylalanine Levels (μmol/L) |  |  |  |  |  |
| Concurrent level | 572 (383) | 113 - 1750 | - | - | - |

*Note:* FSIQ = estimated full scale IQ using the Wechsler Abbreviated Scale of Intelligence- 2<sup>nd</sup> Edition.

All participants with early-treated PKU were diagnosed with classic PKU at birth. [Note that PKU categorization is based on blood Phe levels at the time of diagnosis (Classic: >1200 μmol/L; Moderate: 900–1200 μmol/L; Mild: 600–900 μmol/L; Hyperphenylalaninemia: 120–600 μmol/L).] All PKU participants began a Phe-restricted diet shortly thereafter, as indicated by medical records and/or patient report. Seven participants were prescribed sapropterin, and five participants were prescribed pegvaliase at the time of testing. Participants with early-treated PKU were recruited from metabolic clinics across the country. Individuals were excluded if they had a history of major neurologic condition unrelated to PKU (e.g., severe head injury). Individuals in the non-PKU group were recruited from the general community.

### 2.2 Materials and Procedure

This study was approved by the University of Missouri Internal Review Board (Review ID# 2092230) and adhered to the principles outlined in the World Medical Association Declaration of Helsinki. Prior to participation, informed consent was obtained from all individuals.

All aspects of participant enrollment and data collection were conducted remotely. EMA data on momentary anxiety and depressive symptoms were collected four times daily (approximately every three hours) over seven consecutive days. Retrospective mood symptom data (i.e., participants were asked to report how they had felt over the previous week) were collected on the day following the seven-day EMA period. EMA data were obtained using a secure smartphone application (TigerAware; Morrison et al., 2018), which participants downloaded to their personal mobile devices after receiving instructions on its use. In addition, participants received prompts at each EMA time point to complete a brief (approximately three-minute) web-based processing speed task.

### 2.3. Anxiety and Depression Assessment

Anxiety and depressive symptoms were assessed using a modified version of the National Institutes of Health (NIH) Patient-Reported Outcomes Measurement Information System (PROMIS) Adult 8-Item Depression and 7-Item Anxiety Short Form instruments, respectively (Bjorner et al., 2014; Pilkonis et al., 2011). Because the PROMIS measures were originally designed to assess symptoms retrospectively over the previous seven days, item wording was slightly modified to reflect the momentary nature of EMA reporting. For example, the item “In the past 7 days, I felt fearful” was adapted to “Right now, I feel fearful.” The 5-point Likert response options were similarly modified, such that “Never, Rarely, Sometimes, Often, and Always” became “Not at all, Only a little, Somewhat, Rather much, and Very much.” The original PROMIS item wording and response scales were retained for the retrospective mood assessment administered on Day 8.

### 2.4. Processing Speed Task

The TestMyBrain Digit Symbol Matching task was used to assess processing speed performance. TestMyBrain.org is a web-based software platform for collecting performance data on cognitive tasks such as simple speeded response tasks. The task is adapted from the Digit Symbol Coding subtest of the Weschler intelligence scales. In brief, participants are shown a key pairing nine abstract symbols with specific digits, which remains visible throughout the task. Participants are presented with a series of symbols one at a time. For each symbol, they must respond with the correct matching digit (based on the aforementioned key) as quickly as possible. The total number of correct responses in 90 seconds is recorded, along with error rate and response times.

### 2.5. Blood Phe Levels

Concurrent blood Phe levels were measured directly on Days 1, 4, and 7 of the study using dried blood spot filter paper testing. Self-reported data on historic metabolic control was also collected. Participants were asked to report their average blood Phe levels during childhood (0–11 years), adolescence (12–17 years), and adulthood (18+ years), as well as the most recent blood Phe level they could recall. Response options were categorized as 0-360, 360-600, 600-1200, and >1200 μmol/L.

### 2.6. Data Analysis Approach

All analyses were conducted in R version 4.4.0 (R Core Team, 2024). Descriptive statistics for demographic and EMA data were calculated (see Tables 1 and 2). For categorical demographic variables, χ^2^ tests were used to test group differences between PKU and non-PKU. Independent samples *t*-tests were used to test for PKU and non-PKU differences in continuous demographic variables, aggregate and retrospective levels of anxiety and depression symptoms, and processing speed variables. Correlations of mood and processing speed measure variables (person-averages across all moments) with average concurrent and self-reported historic Phe levels were completed using either Pearson correlations or Spearman rank-order correlations as appropriate. To help prevent Type I errors and correct for multiple comparisons, a false discovery rate (FDR; *q* < 0.05; Benjamini & Hochberg, 1995) was used with the correlation analyses.

**TABLE 2.** Aggregate ecological momentary assessment (EMA) and retrospective group differences.

| Variable | PKU |  | Non-PKU |  | Group Difference |  |  |
| --- | --- | --- | --- | --- | --- | --- | --- |
|  | *M* | *SD* | *M* | *SD* | *t*<sup>a</sup> | *p* | *g* |
| Aggregate EMA |  |  |  |  |  |  |  |
| Anxiety score | 10.01 | 3.2 | 8.48 | 1.5 | 2.30 | 0.03* | .60 |
| Depression score | 9.9 | 3.1 | 8.56 | 0.7 | 2.24 | 0.03* | .58 |
| PS score | 27.8 | 2.3 | 29.2 | 2.5 | -2.06 | 0.04* | -.54 |
| PS proportion correct | 0.98 | .02 | .98 | .02 | -0.21 | 0.83 | -.06 |
| PS median response time | 693 | 86 | 649 | 71 | 2.11 | 0.04* | .55 |
| PS response time SD | 164 | 69 | 170 | 57 | -0.33 | 0.74 | -.09 |
| Retrospective Mood |  |  |  |  |  |  |  |
| Anxiety Score | 13.67 | 5.1 | 12.42 | 3.7 | 0.99 | 0.32 | .28 |
| Depression Score | 12.44 | 4.7 | 11.29 | 2.9 | 1.04 | 0.30 | .29 |

*Note:* EMA = Ecological momentary assessment, PKU = Phenylketonuria, PS = Processing speed.
<sup>a</sup>Degrees of Freedom = 54 for aggregate EMA, 49 for retrospective mood.
\* $p < .05$

For EMA measures, null multilevel models (MLMs) for each variable with no covariates were fit to estimate the proportion of variance accounted for by random intercepts, indicating between-person variance, and unexplained residuals, indicating within-person variance, to calculate intraclass correlations (ICCs). Given the primary focus on mood, potentially meaningful group differences in anxiety and depression trajectories based on time-related variables (e.g., overall study time point, day of the week, time point in the day) were then explored. In the final MLMs, an interaction term between the group and the time-related variables, both linear change over time (e.g., change over the course of the study) and categorical (e.g., differences in days of the week), was used to assess group differences in symptom change over time. Time-related variable reference points (i.e., coded 0) were the beginning of the study for overall continuous time point (0-27), Sunday for day of the week (0-6), and the first time point in the day (0-3). The non-PKU group and female sex were also coded as 0. Models were estimated using the lme4 (Bates et al., 2015) and lmerTest packages (Kuznetsova et al., 2017) utilizing restricted maximum likelihood. All MLM models began as maximal models (Barr et al., 2013) given the clustered structure of the data. These initial models included random intercepts and random slopes in models containing continuous time-related variables to model between-person differences in anxiety and depression trajectories. If models did not converge or displayed boundary issues regarding the correlation between random intercepts and slopes when applicable, random intercepts and slopes were first modeled without their correlation, followed by removal of random slopes if necessary. Critical alpha level was 0.05 for the fixed effects of PKU group differences based on areas of interest.

Two reliability metrics were calculated for each mood outcome and response time of the processing speed task using Generalizability Theory (GT) equations adapted to the EMA context, utilizing variance components related to the person, time, items, and their three two-way interactions (Shrout & Lane, 2012). Between-person reliability (consistency of measuring individual differences in the assumed underlying construct when aggregating across persons and time points) and within-person reliability (consistency of change or variation in the assumed underlying construct within a subject from one time point to the next) of our data were calculated using the procedure described by Shrout and Lane (2012). In order to estimate how many EMAs were necessary to achieve near maximum reliability based on our original design, between-person reliability estimates (R_KF_) were also calculated for each outcome as a function of the number of overall EMA time points possible, spanning the number of days (one through seven) and number of time points each day (one through four). Within-person reliability calculations (R_C_) do not include the number of time points of collection in their estimation beyond the person-by-time variance component. Confidence intervals were calculated for the reliability estimates of each outcome using Monte Carlo simulation, in which the variance estimates from the initial GT analysis of the full sample were used as inputs. One thousand samples were drawn, in which each of the variance parameters was randomly selected from its own estimated sampling distribution for a given sample, and then individual data were generated according to those values and the specified number of time points per day, total days, and items within subscales. GT analyses were then performed, and reliability estimates were calculated for each sample to construct sampling distributions for each. This was repeated for all permutations of time points per day (1-4), days (1-7), and items (1-7/8/35, for anxiety, depression, and processing speed, respectively).

## 3. RESULTS

Average compliance for the EMA prompts was 82.8% across the sample, with no significant difference between the PKU group (*M* = 81.2%) and the non-PKU group [*M* = 84.3%; *t*(58) < 1, *p* = .484, *g* = .18). Four participants (2 PKU, 2 Non-PKU) completed fewer than 60% of administered prompts and thus were excluded from further analysis. Five participants (one PKU, 4 Non-PKU) did not complete the retrospective symptom measures.

### 3.1. Anxiety & Depression Symptoms

As detailed in Table 2, the PKU group reported significantly higher anxiety symptoms compared to the non-PKU group as measured by EMA [*M*_PKU_ = 10.0, *M*_NonPKU_ = 8.5; *t*(54) = 2.30, *p* = .025, *g* = .60]. A similar pattern was observed for EMA depression symptoms [*M*_PKU_ = 9.9, *M*_NonPKU_ = 8.6; *t*(54) = 2.24, *p* = .030, *g* = .58]. In contrast, no significant group differences were observed for the retrospective symptom measurements of anxiety or depression gathered on Day 8 [*t*(49) < 1.05, *p* > .291, *g* < .30 in both instances].

Correlations between symptom measures and Phe levels (as a marker of metabolic control) are included in Table 3. Within the PKU group, there was a trend (albeit non-significant) reflecting a general association between higher anxiety and depression symptom report and higher Phe levels across all developmental epochs (childhood, adolescence, adulthood, and concurrent; *r* = 0.13-0.41). This pattern was observed for both EMA and retrospective measures.

**TABLE 3.** Between-subject correlations of aggregate ecological momentary assessment (EMA) and retrospective measures with historic self-report and concurrent blood Phenylalanine levels.

| Variable | Phenylalanine Levels ($\mu\text{mol/L}$) | | | | |
| --- | --- | --- | --- | --- | --- |
|  | Self-report<br>for<br>childhood<br>(0-11 yrs) | Self-report<br>for<br>adolescence<br>(12-17 yrs) | Self-report<br>for<br>adulthood<br>(18+ yrs) | Self-report<br>for most<br>recent level | Mean<br>concurrent<br>blood level |
| Aggregate EMA |  |  |  |  |  |
| Anxiety score | 0.24 | 0.26 | 0.30 | 0.26 | 0.19 |
| Depression score | 0.27 | 0.26 | 0.13 | 0.30 | 0.36 |
| PS score | -0.33 | -0.29 | 0.00 | 0.06 | -0.01 |
| PS proportion correct | 0.03 | -0.08 | -0.10 | -0.16 | -0.05 |
| PS median response<br>time | 0.34 | 0.15 | -0.02 | -0.06 | 0.00 |
| PS response time SD | -0.02 | 0.26 | -0.10 | -0.13 | -0.17 |
| Retrospective Mood |  |  |  |  |  |
| Anxiety Score | 0.30 | 0.27 | 0.28 | 0.32 | 0.25 |
| Depression Score | 0.15 | 0.30 | 0.33 | 0.41* | 0.28 |

*Note:* EMA = Ecological momentary assessment, PS = Processing speed.
\* $p < .05$ , uncorrected

#### 3.1.1. Between- and Within-Person Variance

Intraclass correlations for the EMA variables are included in Table S1 of the Supplemental Materials. Based on the unconditional MLM, between-person effects accounted for a slightly larger proportion of variance in EMA anxiety symptoms (52%) as compared to within-person effects (48%) in the overall sample. Of note, individuals with PKU exhibited less within-person variance in EMA anxiety scores (44%) compared to individuals in the non-PKU group (68%).

For EMA depressive symptoms, between-person effects (64%) again accounted for more variance than within-person effects (36%) in the overall sample. When comparing across groups, individuals with PKU exhibited markedly less within-person variance in EMA depression scores (29%) compared to individuals in the non-PKU group (84%).

#### 3.1.2. Time-Related Group Differences

There was a significant difference by group in categorical change of depression symptom ratings from Sunday compared to Friday (See Table 4; PKU Group X Friday *B* = -1.10, *SE* = 0.35, *t* = -3.10, *p* = .002). Visualization of depression symptoms across days of the week (see Figure 1) shows that among the PKU group, depression ratings noticeably dipped on Fridays, whereas Friday depression symptoms in the non-PKU were similar levels to those at the beginning of the week. A similar pattern was observed for anxiety symptoms; however, the effect did not reach significance (PKU Group X Friday *B* = -0.82, *SE* = 0.66, *t* = -1.25, *p* = .219). No other depression or anxiety symptom patterns differed by PKU group based on linear or categorical time-related variables such as overall study time point, day of the week, or time point within a day (all *p* > 0.05).

**TABLE 4.** Group differences in depression and anxiety scores by categorical day of the week.

|  | Estimate | CI | *t* | *p* |
| --- | --- | --- | --- | --- |
| Depression |  |  |  |  |
| Intercept | 8.37 | 7.48 – 9.25 | 18.439 | <0.001* |
| PKU Group | 1.83 | 0.58 – 3.09 | 2.86 | 0.006* |
| Mondays | 0.42 | -0.05 – 0.9 | 1.73 | 0.084 |
| Tuesdays | 0.34 | -0.14 – 0.82 | 1.40 | 0.163 |
| Wednesdays | 0.18 | -0.31 – 0.66 | 0.71 | 0.477 |
| Thursdays | -0.05 | -0.54 – 0.43 | -0.21 | 0.834 |
| Fridays | 0.31 | -0.18 – 0.8 | 1.22 | 0.223 |
| Saturday | 0.17 | -0.32 – 0.65 | 0.66 | 0.509 |
| PKU Group X Mondays | -0.30 | -0.98 – 0.38 | -0.85 | 0.393 |
| PKU Group X Tuesdays | -0.56 | -1.24 – 0.12 | -1.61 | 0.108 |
| PKU Group X Wednesdays | -0.59 | -1.27 – 0.09 | -1.69 | 0.092 |
| PKU Group X Thursdays | -0.29 | -0.97 – 0.4 | -0.81 | 0.416 |
| PKU Group X Fridays | -1.10 | -1.79 – -0.41 | -3.10 | 0.002* |
| PKU Group X Saturdays | -0.64 | -1.34 – 0.06 | -1.79 | 0.075 |
| Anxiety |  |  |  |  |
| Intercept | 8.15 | 7.09 – 9.22 | 14.76 | <0.001* |
| PKU Group | 1.56 | 0.06 – 3.06 | 2.00 | 0.050 |
| Mondays | 0.40 | -0.53 – 1.33 | 0.82 | 0.415 |
| Tuesdays | 0.29 | -0.49 – 1.07 | 0.73 | 0.470 |
| Wednesdays | 0.58 | -0.21 – 1.37 | 1.41 | 0.165 |
| Thursdays | 0.35 | -0.6 – 1.29 | 0.71 | 0.480 |
| Fridays | 0.44 | -0.46 – 1.33 | 0.94 | 0.353 |
| Saturday | 0.18 | -0.5 – 0.87 | 0.52 | 0.603 |
| PKU Group X Mondays | 0.31 | -1.02 – 1.63 | 0.45 | 0.658 |
| PKU Group X Tuesdays | 0.11 | -1 – 1.21 | 0.19 | 0.851 |
| PKU Group X Wednesdays | 0.07 | -1.05 – 1.18 | 0.12 | 0.907 |
| PKU Group X Thursdays | 0.52 | -0.81 – 1.85 | 0.75 | 0.456 |
| PKU Group X Fridays | -0.82 | -2.08 – 0.44 | -1.25 | 0.219 |
| PKU Group X Saturdays | -0.42 | -1.4 – 0.56 | -0.83 | 0.409 |

*Note.* PKU = Phenylketonuria, CI = Confidence interval. Sunday was coded as 0.
\* $p < .05$

**Figure 1.**
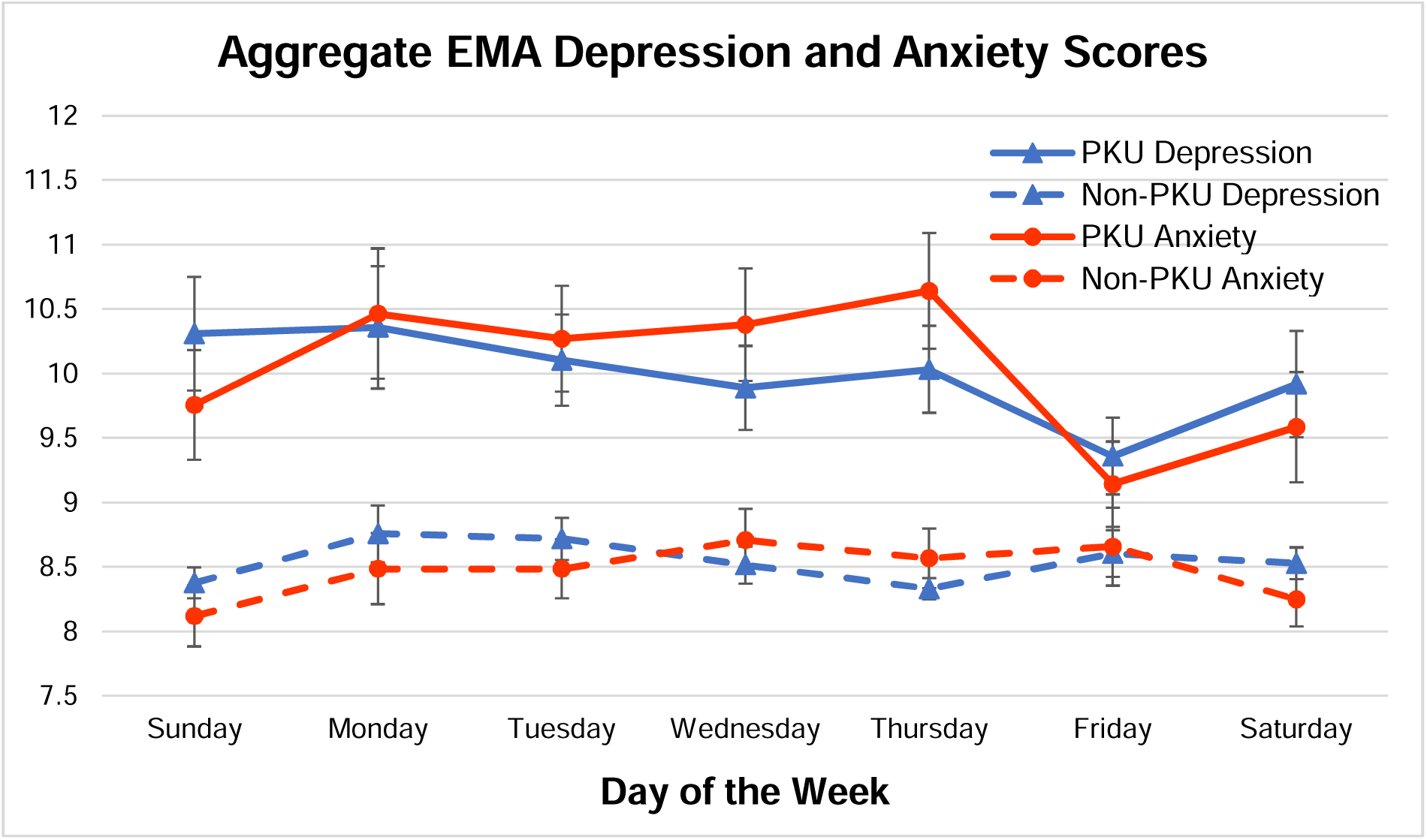
Depression scores by day of the week for individuals with and without PKU. *Note*: PKU = Phenylketonuria, EMA = Ecological momentary assessment.

### 3.2. Processing Speed Performance

The PKU group performed more poorly than the non-PKU group on the processing speed task as reflected by both fewer total number of correct responses (*M*_PKU_ = 27.8, *M*_Non-PKU_ = 29.2) and slower median response time (*M*_PKU_ = 693 ms, *M*_Non-PKU_ = 649 ms) [*t*(54) > 2.10, *p* < .05, *g* > .53 in both instances]. As evident in Table 2, the groups performed comparably in terms of proportion correct and response time variability (*p* > .73 in both instances). Within the PKU group, no relationship was observed between Phe levels–historic or concurrent– and any of the task performance variables (|*r*| < .35, *p* > .092 uncorrected, in all instances).

As detailed in Table S1 of the Supplemental Materials, the proportion of variance accounted for by within-person effects ranged from 47% to 84% for the processing speed outcome variables. The proportion of variance accounted for by between-person effects was comparably low, particularly for and proportion correct (22%) and response time variability (16%). Proportions of within- and between-person variance were generally the comparable in both the PKU and non-PKU groups for each of the processing speed performance variables.

### 3.3. Reliability of EMA Measures

As detailed in Table S2 of the Supplemental Materials, excellent between-person reliability (>0.99) was found over all possible time points for the EMA-adapted PROMIS anxiety and depression instruments, as well as for the processing speed task. Within-person reliability of change over all time points was also acceptable (moderate to good; Shrout, 1998) but varied more across instruments (PROMIS Anxiety scale = 0.85; PROMIS Depression scale = 0.81, Processing Speed task = 0.74).

Between-person reliability as a function of the completed number of EMAs per the original study design is shown in Figure 2. For all instruments, maximum between-person reliability was observed after approximately 10 EMA completions within the context of our study design. Overall, there were no significant differences in reliability between PKU and non-PKU groups (supplemental Table S2), although individuals with PKU had slightly higher within-person reliabilities in mood scores and lower within-person reliability for processing speed response time.

**Figure 2.**
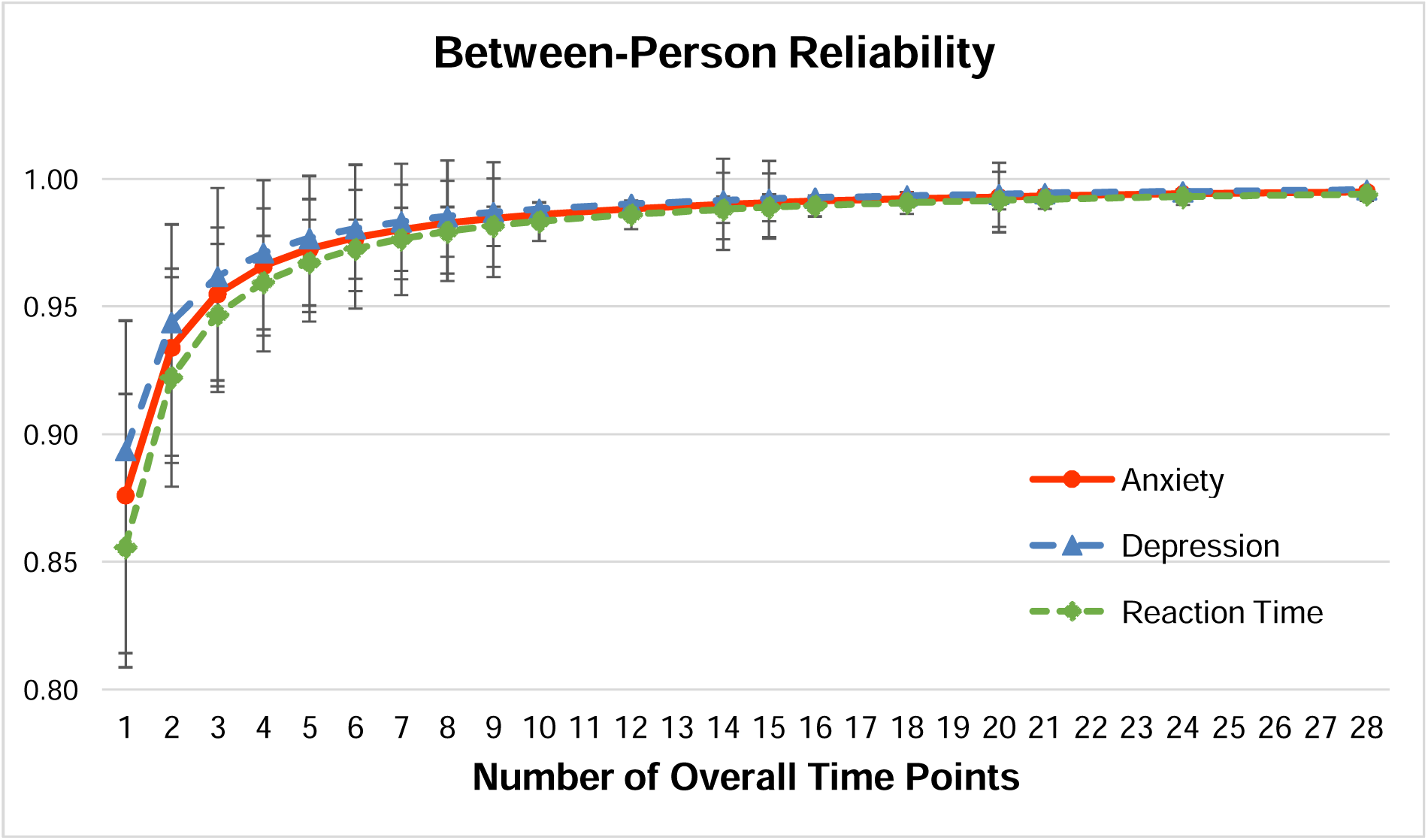
Between-person reliability as a function of number of EMAs completed based on design by outcome. *Note*: EMA = Ecological momentary assessment.

## 4. DISCUSSION

While several studies have reported elevated symptoms of anxiety and depression in individuals with early-treated PKU (e.g., Boland et al., 2024; Bilder et al., 2013; Koch et al., 2002; Manti et al., 2016), others have not found evidence of increased mood-related symptoms in this population (e.g, Brumm et al., 2004; Channon et al., 2007; Clacy et al., 2014; Palermo et al., 2020). The current study sought to use EMA, a potentially more sensitive and ecologically valid method than traditional retrospective report measures, to further examine the impact of early-treated PKU on anxiety and depressive symptoms.

In the present study, individuals with early-treated PKU reported higher levels of anxiety and depression across EMA assessment occasions than individuals without PKU. As discussed previously, findings regarding mood symptoms in early-treated PKU have been mixed. Several studies have reported elevated rates of anxiety and depression symptoms among individuals with early-treated PKU (e.g., Boland et al., 2024; Bilder et al., 2013; Waisbren et al., 2017), whereas others have not observed this pattern (e.g., Brumm et al., 2004; Channon et al., 2005, 2007). The elevated mood symptoms observed in the present study likely reflect the combined influence of neurobiological and psychosocial factors. Dysregulation of neurotransmitter systems, including dopamine, serotonin, and norepinephrine, which play critical roles in emotional regulation, may contribute to increased symptoms of anxiety and depression (e.g., Hasler, 2010; Manti et al., 2016; ten Hoedt et al., 2011). From a psychosocial perspective, the ongoing demands associated with managing a chronic condition such as PKU may represent a significant source of stress, thereby increasing vulnerability to anxiety and depressive symptoms (Bodner et al., 2024; Stanton et al., 2007).

In contrast to the EMA findings, retrospective measurements of mood showed no significant group differences. This discrepancy could be illustrative of elements from past contrasting findings (Channon et al., 2005; Clacy et al., 2014; Palermo et al., 2020), and suggest that appropriately comprehensive EMA measurement may be more sensitive than traditional approaches which rely on a single-time retrospective assessment. Potential factors contributing to EMA’s increased sensitivity include real-time data collection, large data sampling, and its inherent limiting of challenges associated with memory and/or introspection difficulties. However, additional research is warranted to replicate the reported differences.

The PKU group was also found to show slower response times and lower scores as compared to the non-PKU group on the EMA digit symbol matching task. This finding supports commonly reported processing speed deficits in those with early-treated PKU (Christ et al., 2010; Clocksin et al., 2023). The current study’s demonstrated adequate reliability of processing speed measurement through repeated digital assessment within EMA is particularly encouraging in its benefit to assessing everyday cognitive functioning, including over time, in rare clinical samples such as PKU (Singh et al., 2021, 2023).

Contrary to some prior studies (Clacy et al., 2014; Clocksin et al., 2023; Didycz & Bik-Multanowski, 2018), we found no relationship between metabolic control (as reflected by Phe levels) and mood symptom severity or processing speed in the PKU group when correcting for multiple comparisons. As discussed above, it may be that PKU is generally associated with an increased risk of anxiety and depression, possibly related to the stress associated with management of a chronic illness (Manti et al., 2016). In addition, a limitation of the present study is that minimal information on historical treatment history was available for participants with PKU. Despite efforts to obtain comprehensive treatment records, data collection was constrained by common challenges in adult PKU populations, including transitions between healthcare providers, unavailable or destroyed historical records, and periods of loss to clinical follow-up. Although no association was observed between concurrent blood Phe levels and mood symptoms, mood outcomes may be influenced by longer-term metabolic control (e.g., over the preceding year) or Phe exposure during earlier developmental periods (e.g., adolescence). These relationships may not have been detectable in the current study because of aforementioned limitations in available historical data. Further research is needed to clarify the associations between metabolic control and mood symptomatology.

Multilevel modeling revealed few significant time-related group differences, indicating broadly similar temporal patterns of anxiety and depression symptoms in individuals with and without PKU across the study period, weekdays, and daily time points. However, symptom levels were generally higher in the PKU group. Notably, individuals with PKU reported significantly lower depression symptoms on Fridays compared to Sundays, whereas no significant weekday differences were observed in the non-PKU group. A similar, though nonsignificant, pattern was observed for anxiety. The nature of this finding remains unclear. The Friday reduction in mood symptoms among individuals with PKU may reflect meaningful real-world influences, such as anticipation of the weekend, employment-related factors, or treatment-management behaviors that vary by weekday. Further research is needed to replicate this finding and clarify the underlying mechanisms.

Using generalizability theory within an EMA framework (Shrout & Lane, 2012), the PROMIS anxiety and depression scales and the TMB digit-symbol matching task demonstrated excellent between-person reliability when aggregated across measurement occasions, consistent with the increased reliability expected from repeated assessments. Within-person reliability was also adequate for all measures, supporting their use in examining temporal variability in anxiety, depression, and processing speed. Similar to previous reports of cognitive task reliability (Sliwinski et al., 2018), the processing speed measure showed lower within-person reliability than the mood scales, although reliability remained acceptable. Reliability estimates indicated that relatively few EMA assessments were required to achieve near-maximal between-person reliability, with minimal gains beyond 10 timepoints. However, these estimates should be interpreted in light of the study’s specific design, assessment schedule, and sample characteristics.

### 4.1. Additional Limitations and Future Directions

Blood Phe concentrations were collected on three occasions during the study period. Future studies incorporating more frequent assessments, potentially on a daily basis, would enable a more detailed examination of the relationship between short-term fluctuations in metabolic control and psychological or cognitive outcomes. Recent advances in home Phe monitoring technologies, which allow for regular testing and immediate feedback, may facilitate such investigations (Wada et al., 2023).

The reliability findings suggest that the measures used provide reliable estimates of aggregate anxiety, depression, and processing speed when scores are averaged across repeated assessments. However, caution is warranted when interpreting scores from a single EMA occasion as indicators of stable individual characteristics, given the proportion of momentary, within-person variability ranging between 35-80% across measures. Replication in larger samples of early-treated individuals with PKU is needed, along with further exploration of EMA data. Larger samples would also permit more robust examination of within-person associations and day-to-day changes, rather than primarily focusing on group-level differences (e.g., Kratz et al., 2017).

Overall, the present study supports the sensitivity and reliability of EMA measures for detecting PKU-related differences in mood and processing speed within a cross-sectional framework. Future research should evaluate the responsiveness of these measures to longitudinal changes in metabolic control, including reductions in Phe concentrations and improvements in dietary management (e.g., increased natural protein intake) associated with emerging treatment approaches.

### 4.2. Summary & Conclusions

Using ecological momentary assessment (EMA), we found that individuals with early-treated phenylketonuria (PKU) reported greater symptoms of anxiety and depression and demonstrated poorer processing speed performance compared with a non-PKU comparison group. Importantly, these group differences were not detected using traditional retrospective self-report measures administered at a single time point. These findings underscore the value and feasibility of EMA for advancing understanding of psychological symptoms and neurocognitive functioning in individuals with early-treated PKU. By reducing limitations commonly associated with retrospective assessments, including recall bias and inaccuracies in symptom recollection, EMA may provide a more accurate characterization of symptom burden and daily functioning in this population. EMA may be especially well suited for rare disorders such as PKU, in which limited population size often constrains sample recruitment, as intensive longitudinal data collection can enhance statistical power to detect clinically meaningful effects while capturing experiences in naturalistic settings. Future research should examine within-person associations among psychological symptoms, cognitive performance, and physiological factors, as well as evaluate intervention-related changes over time in individuals with PKU.

## DATA AVAILABILITY

The data that support the findings of this study may be available from the corresponding author (Shawn E. Christ) upon reasonable request.

## DECLARATION OF GENERATIVE AI AND AI-ASSISTED TECHNOLOGIES IN THE MANUSCRIPT PREPARATION PROCESS

During the preparation of this work, the authors used Microsoft Copilot to enhance the clarity, organization, and flow of selected sections of the manuscript. After using this tool, the authors reviewed and edited all generated content as needed and take full responsibility for the content of the published article.

## CREDIT AUTHORSHIP CONTRIBUTION STATEMENT

**Braden E. Hayse**: Formal analysis, Investigation, Writing - Original Draft, Visualization. **Sean P. Lane**: Formal analysis, Writing - Review & Editing. **Shifali Singh**: Investigation, Writing - Review & Editing. **Stephanie Sacharow**: Conceptualization, Funding acquisition, Writing - Review & Editing. **Benjamin D. Goodlett**: Conceptualization, Funding acquisition, Writing - Review & Editing. **Shawn E. Christ**: Conceptualization, Funding Acquisition, Methodology, Investigation, Supervision, Writing - Review & Editing, Visualization.

## ACKNOWLEDGMENTS

We extend our gratitude to the participants and their families for their time and participation, such that we can advance our understanding of PKU. We also thank the National PKU Alliance, various regional PKU advocacy groups, and the faculty and staff at numerous metabolic clinics nationwide for their assistance with recruitment and data collection.

