## Supplemental Materials for "Ecological Momentary Assessment of Anxiety and Depression in Individuals with Early-Treated Phenylketonuria"

### Supplementary Materials

**Table S1.**

*Intraclass correlations for mood and processing speed variables.*

| Variable | Overall | | PKU | | Non-PKU | |
|---|---|---|---|---|---|---|
| | Between-person ICC | Within-person ICC | Between-person ICC | Within-person ICC | Between-person ICC | Within-person ICC |
| Anxiety score | 0.52 | 0.48 | 0.56 | 0.44 | 0.32 | 0.68 |
| | [0.41, 0.60] | [0.39, 0.59] | [0.39, 0.67] | [0.33, 0.61] | [0.19, 0.44] | [0.56, 0.81] |
| Depression score | 0.64 | 0.36 | 0.71 | 0.29 | 0.16 | 0.84 |
| | [0.54, 0.72] | [0.28, 0.46] | [0.56, 0.79] | [0.21, 0.44] | [0.07, 0.24] | [0.76, 0.93] |
| PS score | 0.48 | 0.52 | 0.5 | 0.5 | 0.45 | 0.55 |
| | [0.38, 0.57] | [0.43, 0.63] | [0.33, 0.62] | [0.38, 0.65] | [0.29, 0.59] | [0.44, 0.70] |
| PS proportion correct | 0.22 | 0.78 | 0.21 | 0.79 | 0.24 | 0.76 |
| | [0.15, 0.30] | [0.70, 0.85] | [0.11, 0.31] | [0.69, 0.88] | [0.13, 0.35] | [0.65, 0.87] |
| PS median reaction time | 0.53 | 0.47 | 0.54 | 0.46 | 0.47 | 0.53 |
| | [0.42, 0.62] | [0.38, 0.58] | [0.37, 0.67] | [0.34, 0.62] | [0.31, 0.60] | [0.41, 0.68] |
| PS reaction time SD | 0.16 | 0.84 | 0.2 | 0.8 | 0.13 | 0.87 |
| | [0.10, 0.22] | [0.77, 0.90] | [0.10, 0.30] | [0.69, 0.90] | [0.05, 0.21] | [0.80, 0.95] |

*Note*: ICC = Intraclass correlation, PS = Processing speed. 95% confidence intervals are in brackets.

**Table S2.**

*Between- and within-person reliability coefficients for mood and processing speed variables.*

| Variable | Overall | | PKU | | Non-PKU | |
|---|---|---|---|---|---|---|
| | Between-person Reliability | Within-person Reliability | Between-person Reliability | Within-person Reliability | Between-person Reliability | Within-person Reliability |
| Anxiety score | 0.995<br>[0.993, 0.996] | 0.85<br>[0.843, 0.863] | 0.997<br>[0.995, 0.998] | 0.875<br>[0.863, 0.887] | 0.985<br>[0.978, 0.992] | 0.816<br>[0.795, 0.833] |
| Depression score | 0.996<br>[0.994, 0.997] | 0.809<br>[0.797, 0.822] | 0.997<br>[0.996, 0.998] | 0.822<br>[0.805, 0.84] | 0.959<br>[0.946, 0.98] | 0.789<br>[0.77, 0.808] |
| PS reaction time | 0.994<br>[0.992, 0.996] | 0.736<br>[0.712, 0.756] | 0.996<br>[0.995, 0.998] | 0.712<br>[0.692, 0.752] | 0.991<br>[0.989, 0.996] | 0.758<br>[0.735, 0.787] |

*Note*: PS = Processing speed. 95% confidence intervals are in brackets.

**Figure S1.**

*Overall between-person reliability by outcome as a function of number of EMA days completed assuming all four timepoints.*

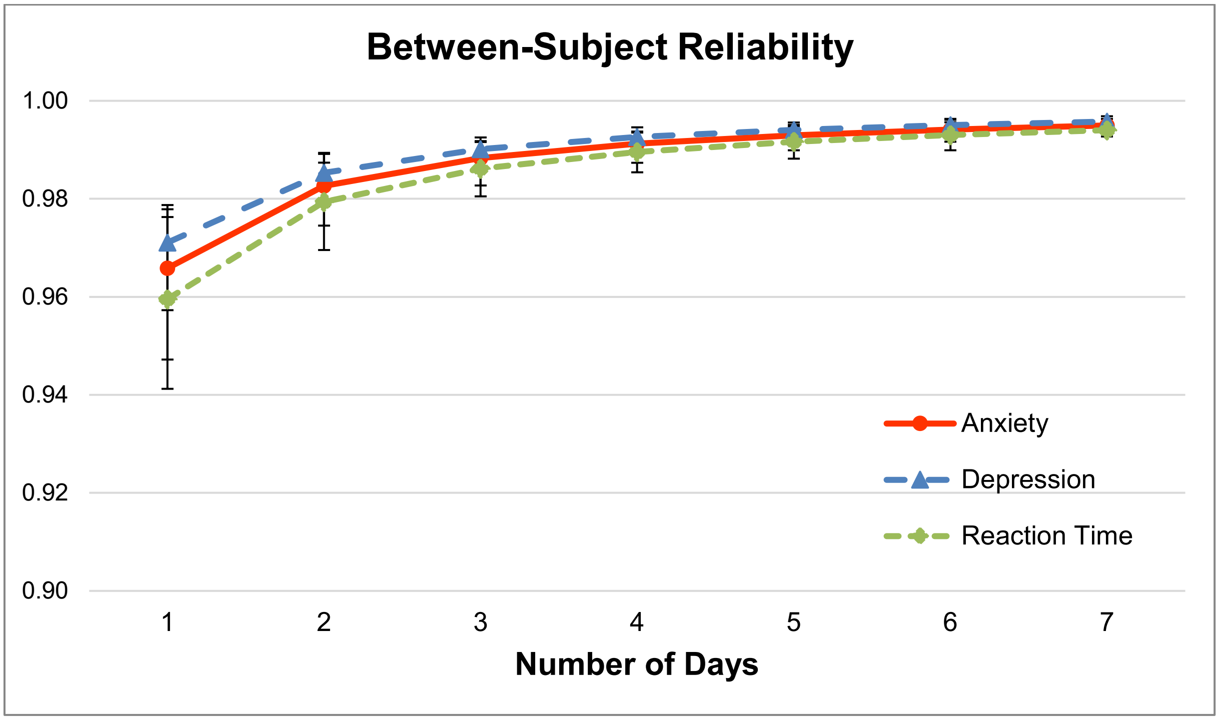


*Note*: EMA = Ecological momentary assessment.

**Figure S2.**

*Overall between-person reliability by outcome as a function of number of EMA daily timepoints completed assuming all seven days.*

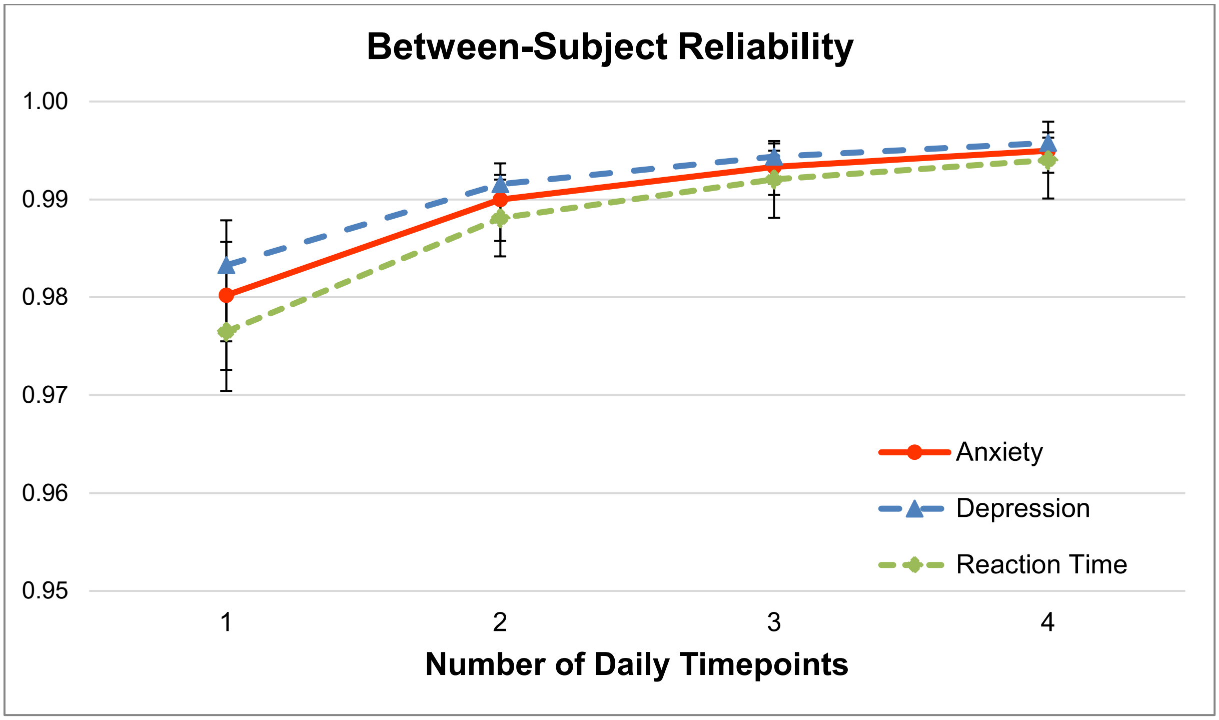


*Note*: EMA = Ecological momentary assessment.
